# Exercise Interventions Improve Balance of Children with Down Syndrome: A Systematic Review

**DOI:** 10.1101/2025.03.30.25324917

**Authors:** Qing Li, Siyu Wang, Congying OuYang

**Author notes:** Author : Qing Li; Doctoral; Research interests: General Sports Training Theory; E- mail Author : Congying OuYang; Doctoral; Research interests: General Sports Training Theory; Author : Siyu Wang; Master; Research interests: Nursing education.

## Abstract

**Objective:** This study systematically investigates the efficacy of exercise interventions in improving balance ability among children with Down syndrome (DS), aiming to establish a robust evidence-based foundation for future research and clinical practice. Through rigorous methodological analysis, the research seeks to inform the development of scientifically grounded, rational, and efficient personalized exercise intervention programs. The findings are expected to provide both theoretical frameworks and practical guidelines for optimizing motor skill rehabilitation strategies, thereby contributing to the enhancement of physical and mental well-being in this vulnerable population. This investigation underscores the critical intersection of evidence-based kinesiology and special needs rehabilitation, offering multidimensional insights for advancing targeted therapeutic approaches in developmental disability management.

**Methods:** A comprehensive literature search was conducted across multiple electronic databases including PubMed, Embase, Web of Science, and Cochrane Library to identify relevant studies investigating exercise interventions for balance improvement in children and adolescents with Down syndrome. Two independent researchers systematically performed study screening and data extraction following predefined inclusion/exclusion criteria. The methodological quality of included studies was assessed using the Cochrane Risk of Bias Tool for randomized controlled trials. Statistical analyses were executed utilizing Review Manager 5.4 (RevMan) and Stata 15.1 software packages, with meta-analytical procedures implemented to quantify intervention effects, assess heterogeneity (I² statistics), and evaluate publication bias through funnel plot analysis and Egger’s regression test. This dual-analyst approach with cross-verification mechanisms ensured methodological rigor in data synthesis and interpretation.

**Results:** This systematic review incorporated 17 eligible studies involving 500 children and adolescents with Down syndrome (aged 0-17 years) from multinational cohorts spanning Egypt, India, Germany, Turkey, and other regions. The implemented interventions encompassed core strength training, dual-task balance exercises, traditional Indian dance, and equine-assisted therapy, with outcomes assessed through diverse measurement instruments.Meta-analytic synthesis revealed statistically significant improvements in balance control measured by the Biodex Stability System (BSS) across 7 studies [*I²=74%, pooled SMD=-0.342 (95% CI: -0.450 to -0.233), P<0.01*]. Sensitivity analysis excluding studies with substantial heterogeneity maintained significance [*I²=65%, SMD=-0.387 (95% CI: -0.508 to -0.265), P<0.01*]. Three studies utilizing the Bruininks-Oseretsky Test of Motor Proficiency (BOTMP) demonstrated clinically meaningful enhancements [*I²=62%, MD=8.296 (5.792-10.800), P<0.01*], with heterogeneity fully resolved upon outlier removal [I²=0%, MD=6.868 (4.586-9.150), P<0.01]. Conversely, no statistically significant effects were observed in three trials employing the Pediatric Balance Scale (PBS) [*I²=80%, MD=1.363 (-0.963-3.689), P=0.251*], potentially attributable to measurement insensitivity or intervention-protocol mismatch.

**Conclusion:** The available evidence substantiates that exercise interventions demonstrate significant efficacy in enhancing postural control and balance performance among children and adolescents with Down syndrome.

---

Down syndrome (DS), a neurogenetic disorder caused by trisomy 21, is pathophysiologically characterized by multisystem manifestations including ligamentous laxity, generalized hypotonia, osteopenia, intellectual disability, and delayed sensorimotor integration. These constitutional impairments collectively contribute to compromised postural control mechanisms, resulting in developmental delays in motor skill acquisition and persistent challenges in dynamic balance regulation throughout the lifespan[1].Evidence from developmental kinesiology research indicates that children with Down syndrome exhibit significant deficits in fundamental movement skills compared to neurotypical peers[2], particularly in postural control mechanisms[3][4]. Balance competence, defined as the neurophysiological capacity to maintain gravitational alignment during static and dynamic tasks, serves as a critical determinant of functional independence in daily living[5]. This multidimensional construct not only supports physical health through enhanced musculoskeletal coordination but is also closely intertwined with psychological well-being, social interaction capabilities, and cognitive development trajectories. From a biomechanical perspective, postural equilibrium requires the integrated processing of somatosensory inputs (vestibular, proprioceptive, and visual systems), neuromuscular activation patterns, and adaptive weight-shifting strategies to counteract destabilizing forces and prevent falls[6].Current empirical evidence and clinical observations substantiate that children with Down syndrome manifest clinically significant postural instability compared to age-matched typically developing peers, a multifactorial limitation that exacerbates challenges in social participation and attainment of functional independence in activities of daily living (ADL). This functional impairment creates a cyclical pattern of activity restriction that necessitates urgent implementation of targeted neuromotor interventions. The critical role of balance proficiency as a keystone developmental competency, coupled with its profound impact on quality of life metrics in this population, underscores the scientific imperative to establish evidence-based rehabilitation protocols. While structured exercise regimens grounded in motor learning paradigms have demonstrated robust efficacy in enhancing postural adaptability among neurotypical children, the translation of these interventions to the Down syndrome cohort requires systematic adaptation accounting for their unique neuromotor phenotype and sensorimotor processing characteristics[7][8].Emerging advances in the elucidation of pathophysiological mechanisms underlying Down syndrome, concomitant with paradigm shifts in exercise science, have catalyzed systematic investigation into neuromotor rehabilitation strategies. Contemporary research has elucidated the therapeutic potential of targeted exercise modalities in ameliorating sensorimotor deficits, particularly postural control impairments, within this population. This evolving scientific landscape has precipitated focused inquiry into dose-response relationships of various intervention protocols, including Neuromuscular activation training[9] hippotherapy[10] traditional Indian Dance[11]and specific training for core strength and balance ability[13].

Notwithstanding the accumulating evidence exploring exercise-mediated improvements in balance function among children with Down syndrome (DS), substantial methodological heterogeneity across studies—manifested in divergent experimental designs, limited sample sizes, heterogeneous intervention protocols, and psychometrically distinct assessment tools— has engendered inconsistent findings and precluded definitive clinical consensus. While a subset of trials demonstrates statistically significant improvements in postural stability indices, contradictory evidence persists regarding intervention durability and effect magnitude, with particular contention surrounding optimal exercise modalities and dosage parameters. This epistemological ambiguity underscores the exigency for conducting a comprehensive systematic review with meta-analytical synthesis to: (a) quantify pooled effect estimates across heterogeneous study designs; (b) elucidate potential moderators influencing intervention efficacy through subgroup analyses; and (c) establish evidence-based recommendations for clinical translation.The present investigation seeks to address this critical knowledge gap by executing a rigorous evidence synthesis that integrates multi-dimensional outcome metrics (biomechanical, functional, and quality-of-life measures). Through systematic evaluation of dose-response relationships and intervention fidelity factors, this meta-analysis aims to delineate the neurorehabilitative potential of exercise paradigms while identifying key determinants of therapeutic success. The resultant framework will inform the optimization of precision rehabilitation protocols, thereby advancing evidence-based practice guidelines for enhancing postural control competencies and ultimately improving health equity in this neurodevelopmentally vulnerable population.

## 1 Methods

### 1.1 PICOS

The present study was conducted in strict accordance with the Preferred Reporting Items for Systematic Reviews and Meta-Analyses (PRISMA) guidelines to evaluate the effectiveness of exercise interventions on balance capacity in children with Down syndrome based on existing literature. The research framework followed the PICO (Population, Intervention, Comparison, Outcome) model as detailed in Table 1. Throughout the investigation process, two independent researchers systematically performed literature retrieval to identify eligible studies, critically evaluated inclusion and exclusion criteria, conducted data extraction, and performed quality assessment using validated methodological tools. This standardized procedure ensured methodological rigor and minimized potential bias in study selection and data interpretation.

**Table 1.** PICOS.

| Population | Intervention | Comparison | Outcome | Study Design |
| --- | --- | --- | --- | --- |
| Health status | Type of intervention | Acceptance versus acceptance of the intervention | Dynamic Balance Indicators | Randomized controlled trials |
| Down syndrome | Neuromuscular training | Comparison before and after the intervention | Static Balance Indicators |  |
| Functional status | Equestrianism | Comparison of different intervention effects | Dynamic-static balance indicators |  |
| Balance disorders | Dance |  |  |  |
| Ages | Intervention protocols |  |  |  |
| 0-18 | Mode of intervention |  |  |  |
|  | Frequency of intervention |  |  |  |
|  | Duration of intervention |  |  |  |

### 1.2 Search Strategy

In accordance with the PRISMA guidelines, we systematically searched PubMed, Embase, Cochrane Library, and Web of Science databases using a validated search strategy that combined Medical Subject Headings (MeSH) terms with free-text keywords. The final electronic search was executed on December 15, 2024, with no temporal or geographical restrictions applied to the included studies.

The identification of studies of interest was performe dusing 4 groups of search terms:

( Group1 ) Adolescents OR Adolescence Adolescents OR Female OR Adolescent Female OR Female Adolescent OR Female Adolescents OR Adolescents OR Male Adolescent OR Male OR Male Adolescent OR Male Adolescents OR Youth OR Youths OR Teens OR Teen OR Teenagers OR Teenager

( Group2 ) Physical Education OR Training OR Physical Training OR Physical Education OR Physical Training OR Human Activities OR Leisure Activities OR Recreation OR Exercise OR Exercises OR Physical Exercise OR Physical Exercises OR Physical Activity OR Physical Activities OR Aerobic Exercise OR Aerobic Exercises OR Isometric Exercises OR Isometric Exercise OR Acute Exercise OR Acute Exercises OR Exercise Training

(Group3)Down syndrome

( Group4 ) balance OR postural control OR postural sway OR postural stability OR postural instability OR postural adaptation OR postural performance OR postural perturbation OR postural strategy OR posture

### 1.3 Search process

A rigorous dual-reviewer process was implemented for study selection and data extraction. Two independent evaluators (1) conducted parallel literature screening, (2) performed blinded data abstraction, and (3) subsequently cross-verified their findings through consensus meetings. Discrepancies in interpretation (occurring in approximately 15% of cases) were resolved through iterative deliberation, with unresolved issues adjudicated by a third independent arbitrator holding expertise in sports epidemiology.

### 1.4 Eligibility Criteria

#### 1.4.1 Inclusion Criteria

(1)Study Design: This systematic review exclusively incorporated randomized controlled trials (RCTs).(2)Participants: Eligible participants were required to be clinically diagnosed with Down syndrome and aged 0–17 years, consistent with the World Health Organization’s age classification for children and adolescents[12].(3)Interventions: Interventions must be exercise-based and specifically aimed at improving balance function in adolescents with Down syndrome. Studies utilizing non-exercise interventions or those not targeting balance enhancement were excluded. No restrictions were imposed regarding intervention settings (4)Outcomes: Included studies were required to report quantitative outcomes related to pediatric balance metrics, including static balance, dynamic balance, or static-dynamic balance.

#### 1.4.2 Exclusion Criteria

(1) Case studies and non-original studies (reviews, theoretical studies); (2) Non English articles not published in peer-reviewed journals; (3) Republished literature; (4) The age range is not within the range of children and adolescents.

## 2 Results

### 2.1 Study Selection

The protocol for this systematic review has been prospectively registered with the International Prospective Register of Systematic Reviews (PROSPERO) under registration CRD42025633811. As this meta-analysis exclusively utilizes published literature data without direct human subject interventions or primary data collection, it did not require additional ethical approval. All incorporated primary studies were approved by their respective institutional ethics review boards and adhered to internationally recognized ethical standards.

### 2.2 Data Extraction And Quality Assessment

#### 2.2.1 Data Extraction

The data extraction protocol systematically captured:(1)Geographical provenance (country-level data);(2)Temporal parameters (publication year);(3)Study population metrics (total sample size) ; (4) Functional capacity classification (intellectual disability levels assessed via standardized tools) ; (5)Methodological framework (experimental vs. observational designs);(6)Cohort characteristics (intervention/control group age distributions with mean ± SD);(7)Efficacy endpoints (primary and secondary outcomes reported).

### 2.2.2 Quality Appraisal

Quality Appraisal: Dual independent evaluation of methodological rigor was conducted by two authors using the Cochrane Risk of Bias Tool . Each study was classified as low risk, high risk, or unclear risk of bias. Given substantial methodological heterogeneity among included studies, a narrative synthesis approach was adopted where quantitative meta-analysis was precluded.

### 2.3 Study Characteristics

Study Characteristics: The 17 included studies originated from Egypt, India, Germany, Iran, and other countries, all employing randomized controlled trial (RCT) designs with a cumulative sample size of 500 participants. Publication years ranged from 2010 to 2024. Detailed characteristics are presented in Table 2.

**Table 2.** Characteristics of Children’s and Adolescents’ Studies Included in the Systematic Review.

| Study | Country | Year | ID Level (M ± SD) |  | Design | Intervention Group |  |  |  |  | Control Group |  |  |  |  |
| --- | --- | --- | --- | --- | --- | --- | --- | --- | --- | --- | --- | --- | --- | --- | --- |
|  |  |  | Intervention Group | Control Group |  | Sample Size | Boys | Girls | Age Range | Mean Age | Sample Size | Boys | Girls | Age Range | Mean Age |
| Mohamed Ahmed Eid et al<sup>[13]</sup> | Egypt | 2015 | 57.06 (2.98)±2.98 | 57.6 (3.08)±3.08 | RCT | 15 | 8 | 7 | 8-10 | 8.93 (0.7) | 15 | 9 | 6 | 8-10 | 9.26 (0.79) |
| Sukriti Gupta et al<sup>[14]</sup> | India | 2010 | 38–49 | 36–52 | RCT | 11 | 6 | 5 | 7-15 | 13.00 (10.00–14.00) | 12 | 8 | 4 | 7-15 | 13.50 (11.25–14.00) |
| Hojat Allah Amini et al<sup>[15]</sup> | Iran | 2016 | NM | NM | RCT | 8 | NM | NM | 8-10 | 8.6+2.4 | 8 | NM | NM | 8-10 | 8.9+3.70 |
| Yelda Kaya et al<sup>[16]</sup> | Germany | 2022 | NM | NM | RCT | 17 | 13 | 4 | 4-14 | 10.12± 3.30 | 17 | 6 | 11 | 4-14 | 8.18 ± 2.74 |
| Mohamed Ahmed Eid et al<sup>[17]</sup> | Egypt | 2017 | 56.46 ± 5.62 | 57.18±4.38 | RCT | 15 | 8 | 7 | NM | 10.26± 0.79 | 16 | 9 | 7 | NM | 10.05 ± 0.68 |
| Sobhy Aly et al<sup>[18]</sup> | Egypt | 2016 | 48.33 ± 6.38 | 50.33±4.70 | RCT | 15 | 11 | 4 | 6-10 | 8.11 ± 1.26 | 15 | 11 | 4 | 6-10 | 8.34 ± 1.07 |
| Fatima Yasin Khan et al<sup>[19]</sup> | Pakistan | 2023 | NM | NM | RCT | 15 | NM | NM | 8-15 | NM | 15 | NM | NM | 8-15 | NM |
| Neslinur Merve Büyükçelik et al<sup>[20]</sup> | Turkey | 2023 | 5.23±1.24(37–67) | 48.71±11.27 (32–68) | RCT | 13 | NM | NM | 8-18 | 12.08±2.56 (8-18) | 14 | NM | NM | 7-17 | NM |
| Reham Saeed Alsakhaw et al<sup>[21]</sup> | Egypt | 2019 | NM | NM | RCT | 15 | NM | NM | NM | NM | 15 | NM | NM | NM | NM |
| Leiton-Muñoz et al<sup>[22]</sup> | Egypt | 2010 | 36-67 | 36-67 | RCT | 15 | 6 | 9 | 10-13 | 10.92 | 15 | 7 | 8 | 10-13 | 11.56 |
| Saeed Ghaeeni et al<sup>[23]</sup> | Iran | 2015 | NM | NM | RCT | 8 | NM | NM | NM | 9.62±1.68 | 8 | NM | NM | NM | 9.87±1.64 |
| A.R. AZAB et al<sup>[24]</sup> | Egypt | 2022 | 58.19 ± 3.56 | 56.13± 4.10 | RCT | 15 | NM | NM | 7-9 | 9.19 ± 0.75 | 16 | NM | NM | 7-9 | 8.60 ± 0.98 |
| Manasa Kolibylu Raghupathy et al<sup>[25]</sup> | India | 2021 | NM | NM | RCT | 18 | 11 | 7 | 6-10 | 7.8 ± 1.3 | 18 | 10 | 8 | 6-10 | 8.4 ± 1.3 |
| Aliaa Goda et al<sup>[26]</sup> | Egypt | 2024 | 60-70 | 60-70 | RCT | 16 | NM | NM | 6-12 | 16 | 16 | NM | NM | 6-12 | NM |
| Alaa AL-Nemr et al<sup>[27]</sup> | Egypt | 2023 | NM | NM | RCT | 20 | 8 | 12 | 8-10 | 8.86 ± 0. 58 | 20 | 6 | 14 | 8-10 | 9.16 ± 0.48 |
| Mohamed Ahmed Eid et al<sup>[28]</sup> | Egypt | 2022 | NM | NM | RCT | 19 | 11 | 8 | 8-12 | 9.5 ± 1.14 | 19 | 12 | 7 | 8-12 | 9.72 ± 1.36 |
| Samia A et al<sup>[29]</sup> | Egypt | 2010 | NM | NM | RCT | 13 | 5 | 8 | 2-5 | 4.56±0.44 | 13 | 6 | 7 | 2-5 | 3.92±1.16 |

DS = Down syndrome; ID = intellectual disability; NM = not mentioned; RCT = randomized controlled trial;

**Table 3.** Description of Intervention Programs, Control Conditions, and Balance Measures in Children’s Studies.

| Study | Intervention type |  | Intervention characteristics |  |  | Balance Measure |  |  |
| --- | --- | --- | --- | --- | --- | --- | --- | --- |
|  | Intervention group | Control group | Time (min) | Frequency (weekly) | Duration (weeks) | Type | Assessment Method | Parameter |
| Mohamed Ahmed Eid et al<sup>[13]</sup> | (WBV-training) the whole-body vibration training physical therapy program | Conventional physical therapy program | 60 | 3 | 24 | Static balance (OLS: EC + EO)<br>Dynamic balance | BSS | Static-dynamic balance score |
| Sukriti Gupta et al<sup>[14]</sup> | strength and balance training | regular activities followed at school | NM | 3 | 6 | Static balance (OLS: EC + EO)<br>Dynamic balance | BOTMP | Static-dynamic balance score |
| Hojat Allah Amini<sup>[15]</sup> | backward walking exercise | forward walking | 25 | 2 | 8 | Static balance<br>Dynamic balance | BSS | general balance<br>medial collateral<br>anterior-posterior balance indexes |
| Yelda Kaya<sup>[16]</sup> | hippotherapy, home exercise program | home exercise program | 30 | 1 | 6 | Static balance<br>Dynamic balance | PBS | Pediatric Balance Scale (PBS) |
| Mohamed A. Eid et al<sup>[17]</sup> | isokinetic training and conventional physical therapy program | conventional physical therapy program | 60 (15IT+45PT) | 3 | 12 | Dynamic balance | BSS | Static-dynamic balance score |
| Sobhy Aly et al<sup>[18]</sup> | core stability exercises | Conventional physical therapy program | 45-60 | 3 | 8 | Dynamic balance (DLS:EO) | BBS | Overall, A-P, M-L stability index scores |
| Fatima Yasin Khan<sup>[19]</sup> | manual ankle rocking training balance and core stability training | balance and core stability training | 40-60 | 4 | 8 | Static balance (OLS: EC + EO)<br>Dynamic balance | foot function index to check the foot posture that can it maintain the balance of body and static standing balance test to check the stability | Static-dynamic balance score |
| Neslinur Merve Büyükçelik<sup>[20]</sup> | dual-task balance exercises training | No exercise training | 30 | 2 | 8 | Static balance<br>Dynamic balance<br>postural steadiness | PBS | Single Leg Stance Test<br>the Tandem Stance test<br>the 30 s Sit to Stand test |
| Reham Saeed Alsakhaw<sup>[21]</sup> | treadmill training | core stability training | 60 | 3 | 8 | dynamic balance | Berg balance scale and the Biodex Balance System | Functional balance<br>Overall stability index |
| Leiton-Muñoz<sup>[22]</sup> | Wii-Fit | the traditional physical therapy program | NM | 2 | 6 | Static balance (OLS: EC + EO)<br>Dynamic balance | BOTMP | Static-dynamic balance scored |
| Saeed Ghaeeni et al<sup>[23]</sup> | core Stability Training | No exercise training | 45-60 | 3 | 8 | Static Balance | modified stork test | Static Balance |
| A.R. AZAB<sup>[24]</sup> | trampoline-based stretch-shortening cycle exercises | standard physical therapy | 15 | 2 | 12 | Static balance<br>Dynamic balance | BMS | the anterior/posterior stability index (A/P-SI)<br>medial/lateral stability index (M/L-SI)<br>overall stability index (O-SI) |
| Manasa Kolibylu Raghupathy<sup>[25]</sup> | traditional Indian dances | neuromuscular training | 60 | 3 | 6 | Static balance<br>Dynamic balance | PBS | Static-dynamic balance score |
| Aliaa Goda<sup>[26]</sup> | Brain Gym Exercise | traditional balance program | 60 | 3 |  | Static balance<br>Dynamic balance | PBS | SLS、TUG tests<br>pediatric balance scale |
| Alaa AL-Nemr<sup>[27]</sup> | pilates exercise program、physical therapy program | a specially designed physical therapy program | Pilates exercise program 45<br>physical therapy program 45 | 3 | 12 | Dynamic balance | BBS | Anteroposterior<br>Mediolateral<br>Overall |
| Eid MA et al<sup>[28]</sup> | vitamin D in conjunction with aerobic exercises | conventional physical therapy programm | 45 | 3 | 12 | Static balance<br>Dynamic balance | BBS | the medial-lateral stability index<br>the anterior-posterior stability index<br>the overall stability index<br>the 6-Minute Walk Test |
| Samia A et al<sup>[29]</sup> | weight bearing exercises<br>the traditional physical therapy program | the traditional physical therapy program | 30 | 1 | 6 | Static balance<br>Dynamic balance | BOTMP | Static Balance<br>Dynamic Balance<br>Total Balance |

A-P = anterior-posterior; BOTMP = Bruininks-Oseretsky Test of Motor Proficiency; BBS=Biodex Balance System ; BMS=Biodex Medical Systems; PBS=Pediatric Balance Scale; DLS = double-leg stance; EC = eyes closed; EO = eyes open; M-L = mediolateral; NM = not mentioned; OLS = 1-leg stance. Adaptation and fidelity were not mentioned in any study. This test provided a global balance score that measure both static and dynamic balance. dBiodex Medical Systems, Inc., Shirley, New York.

### 2.4 Statistical analysis

Statistical Analysis: Data were analyzed using Review Manager 5.4 and Stata 15.1 software. Heterogeneity was assessed via I² statistics. Fixed-effect models were applied when studies showed homogeneity (I²<50% with p>0.01), while random-effects models were employed for significant heterogeneity (I²≥50% with p<0.01). Subgroup analyses and sensitivity analyses were conducted to investigate sources of substantial heterogeneity. Descriptive synthesis was performed when heterogeneity sources remained undetermined and results were unstable.

### 2.5 Results of meta-analysis

Among the 17 included studies, 7 utilized the Biodex Stability System (BSS). Pooled analysis demonstrated statistically significant effects [I²=74%, 95% CI -0.342 (-0.450 to -0.233), P<0.01]. Exclusion of Study 17 reduced heterogeneity (I²=65%), though remaining substantial. Random-effects model analysis maintained significance *[I^2^=65%,95%CI=-0.387* ( *-0.508* ) *-0.265* ) *,P<0.01]*, confirming BSS’s validity in assessing adolescent balance across exercise modalities despite residual heterogeneity.Three studies employing the Bruininks-Oseretsky Test of Motor Proficiency (BOTMP) showed significant outcomes *[I²=62%, 95% CI 8.296 (5.792-10.800), P<0.01]*. Post-exclusion of Study 2, heterogeneity resolved (I²=0%), with fixed-effect model confirming robustness *[I2=0%,95%CI=6.868* ( *4.586* ) *9.150*)*,P<0.01]*.Contrastingly, three Pediatric Balance Scale (PBS) studies exhibited marked inconsistency *[I^2^=80%,95%CI=1.363* ( *-0.963* ) *3.689* ) *,P=0.251]*, demonstrating non-significant effects (Table 4).

**Table 4.** Meta-analysis results of different scales.

| Outcome | Study | Heterogeneity test results |  | Effect model | Effect (95%CI) | P |
| --- | --- | --- | --- | --- | --- | --- |
|  |  | P | I<sup>2</sup> |  |  |  |
| Biodex Stability System (BSS) | 7 | <0.01 | 74% | Random-effects model | -0.342 (-0.450, -0.233) | <0.01 |
| Exclusion study17 | 6 | 0.013 | 65% | Random-effects model | -0.387 (-0.508, -0.265) | <0.01 |
| Bruininks Oseretsky Test of Motor Proficiency (BOTMP) | 3 | 0.071 | 62% | Random-effects model | 8.296 (5.792, 10.800) | <0.01 |
| Exclusion study 14 | 2 | 0.846 | 0% | Fixed-effect model | 6.868 (4.586, 9.150) | <0.01 |
| Pediatric Balance Scale (PBS) | 3 | 0.080 | 80% | Random-effects model | 1.363 (-0.963, 3.689) | 0.251 |

#### 2.5.1 Biodex Stability System(BSS)

Seven studies employing the Biodex Stability System (BSS) to assess the effects of exercise modalities on adolescent balance demonstrated significant heterogeneity (I²=73.6%, P<0.01), warranting a random-effects model. Sequential exclusion identified Study 17 as the primary heterogeneity source, reducing I² to 65.4% (P=0.013; *Table 5*). Subgroup analyses stratified by intervention type revealed statistically significant differences between:

a. Muscle strength/coordination training (n=4)
b. Core stability/neurocognitive training (n=3)(P<0.01 between subgroups; Table 4).

**Table 5.**
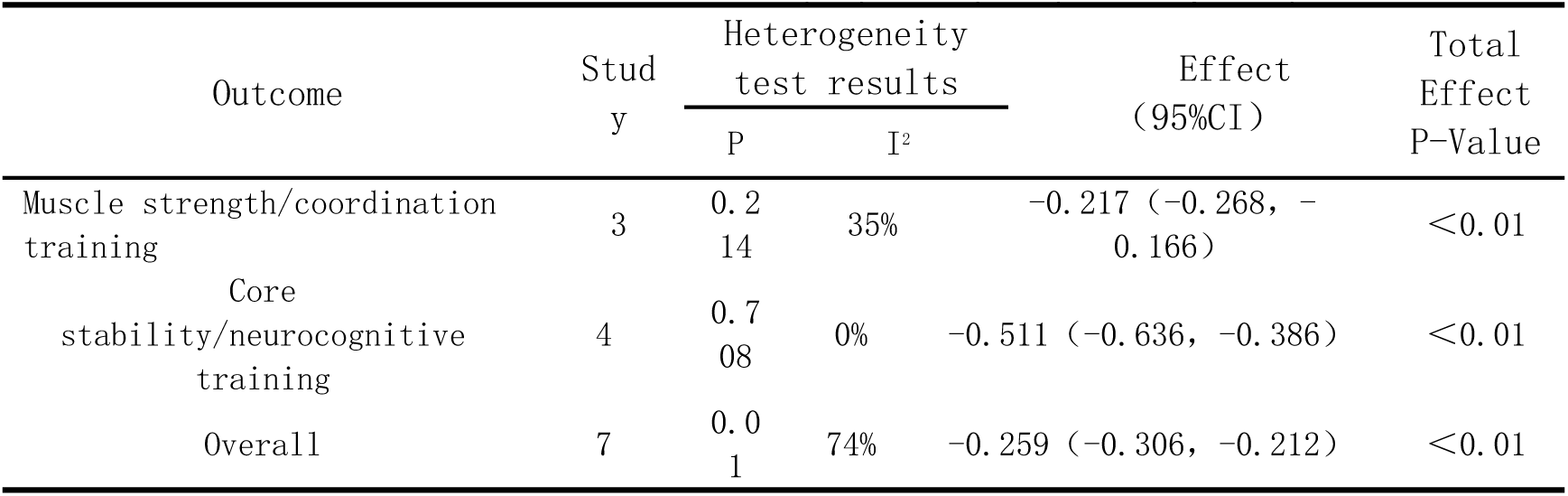
Results of Biodex Stability System (BSS) heterogeneity.

Sensitivity analysis via leave-one-out method confirmed result robustness (Figure 5), with <5% variation in pooled effect size upon individual study exclusion. Publication bias assessment was precluded due to insufficient studies (n<10 per Cochrane criteria).

**Figure 1.**
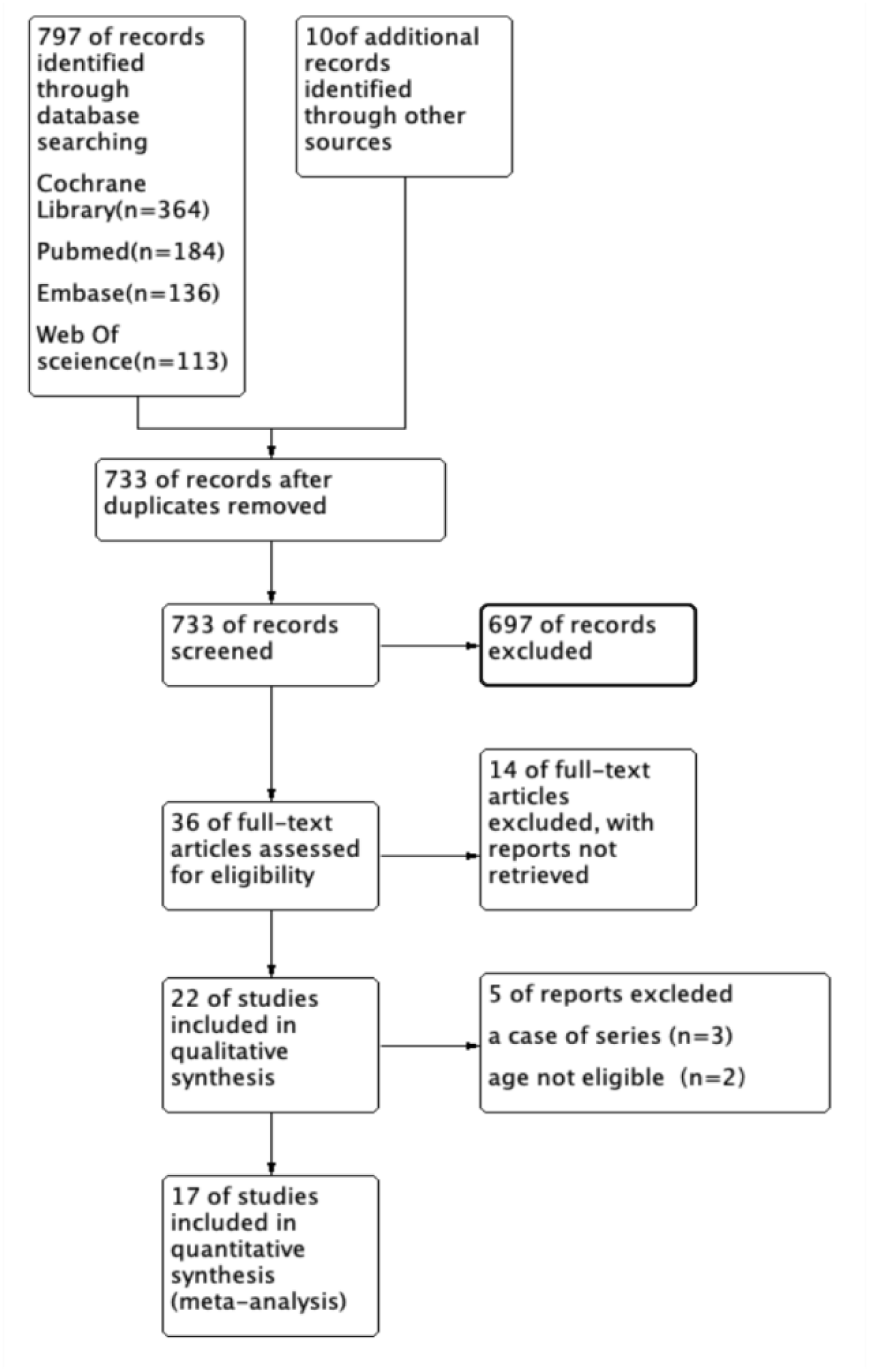
Literature screening flowchart

**Figure 2.**
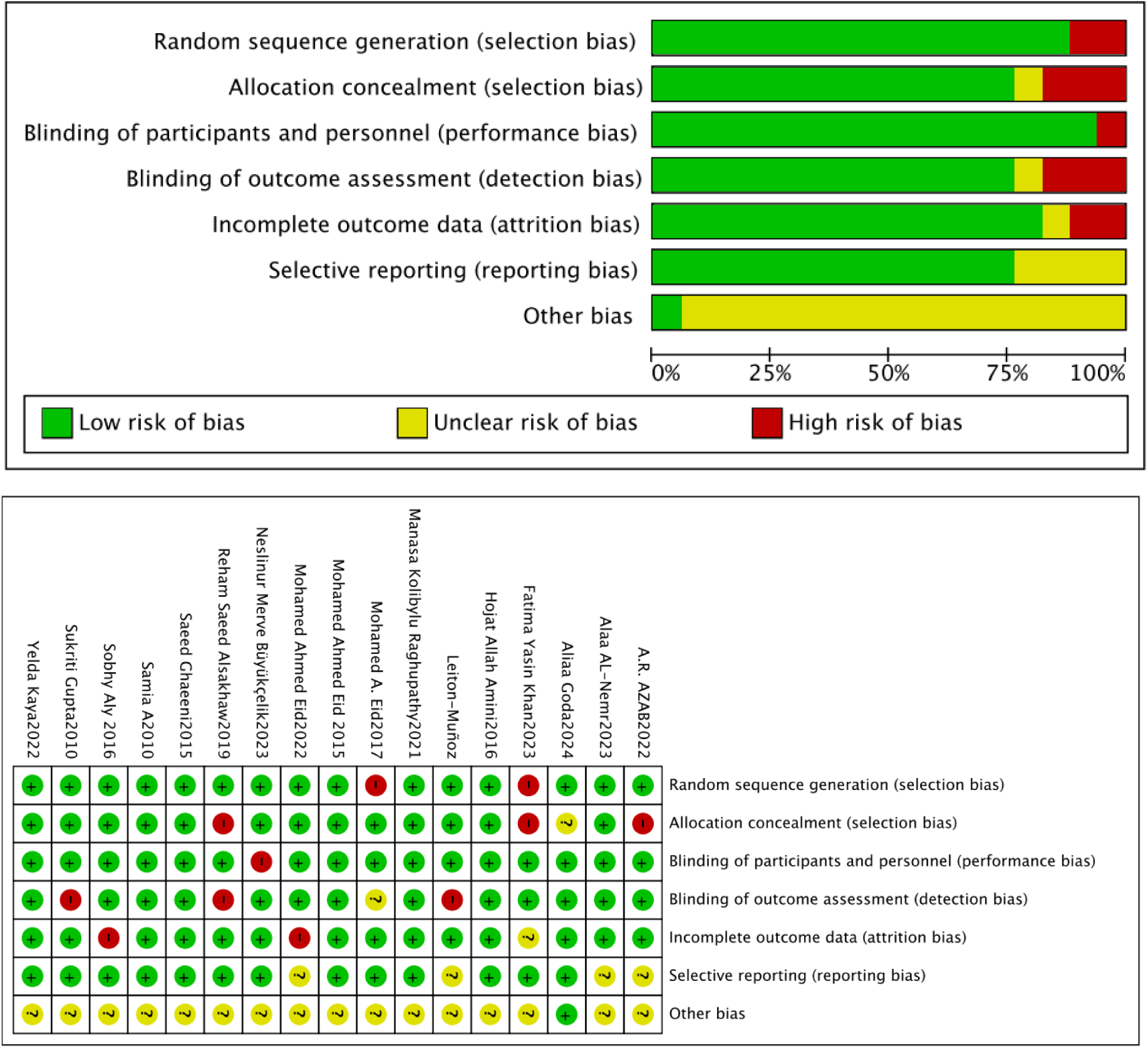
Risk of Bias Assessment Summary

**Figure 3.**
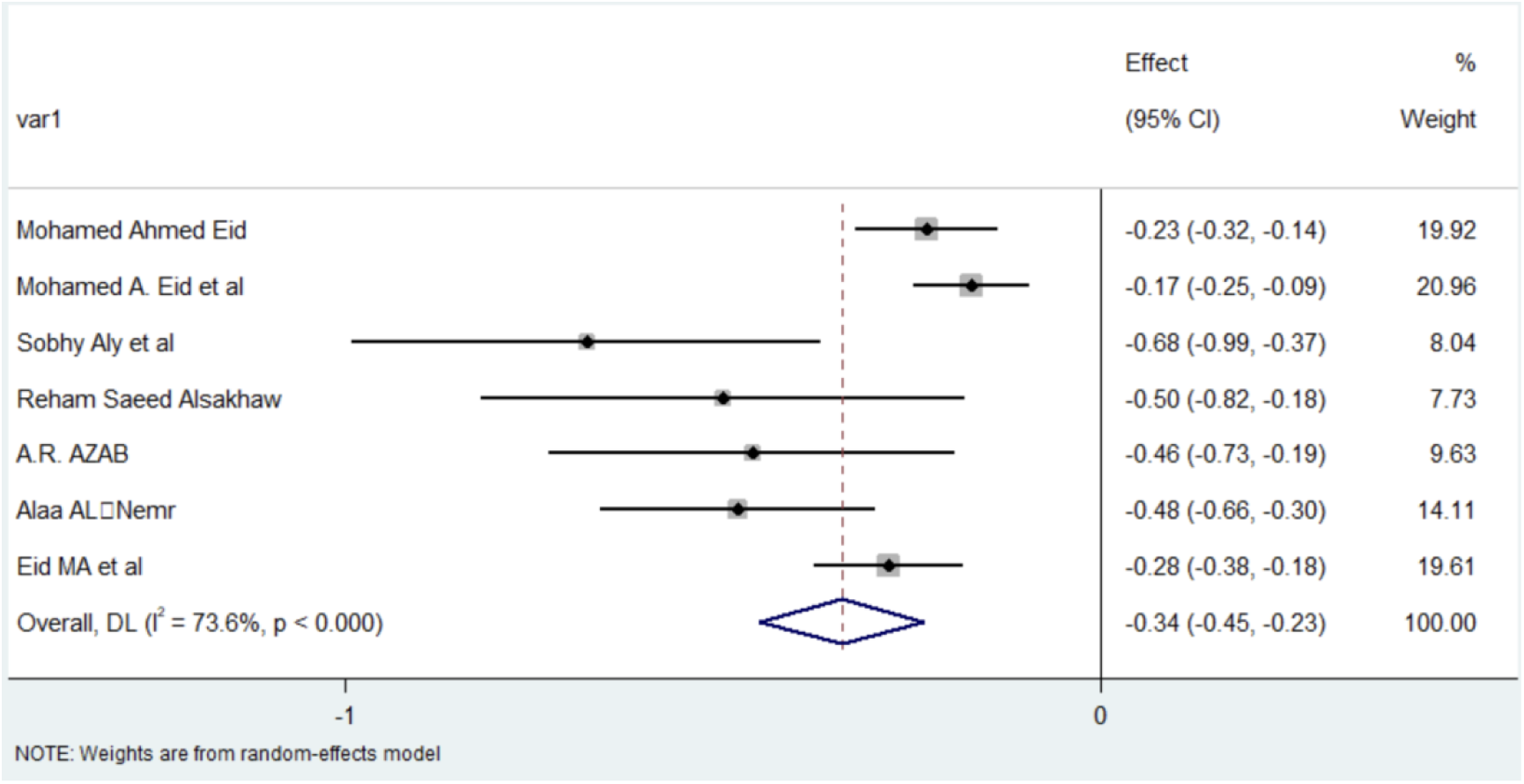
Forest diagram of Biodex Stability System (BSS).

**Figure 3.**
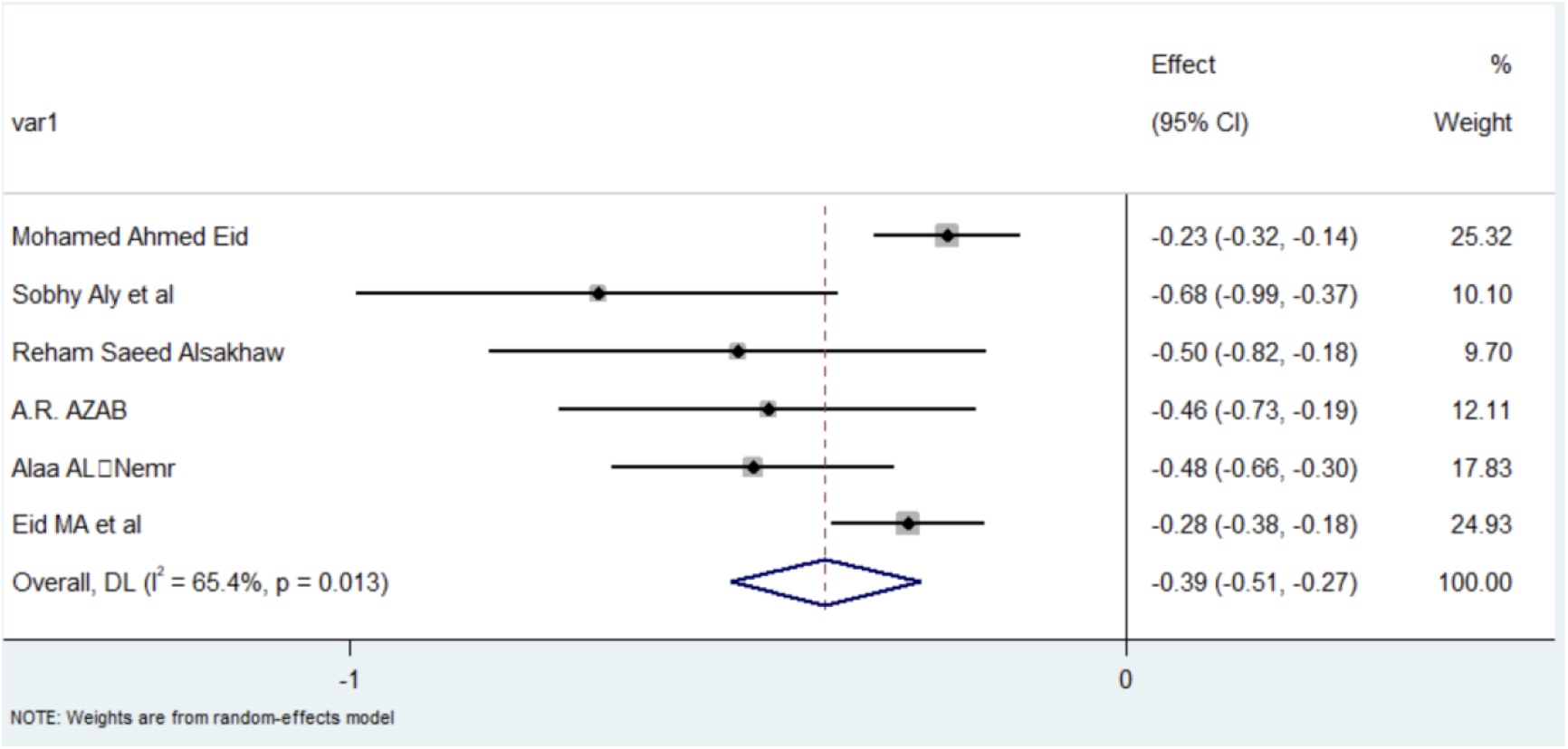
Forest plot after the Biodex Stability System (BSS) exclusion study 2

**Figure 4.**
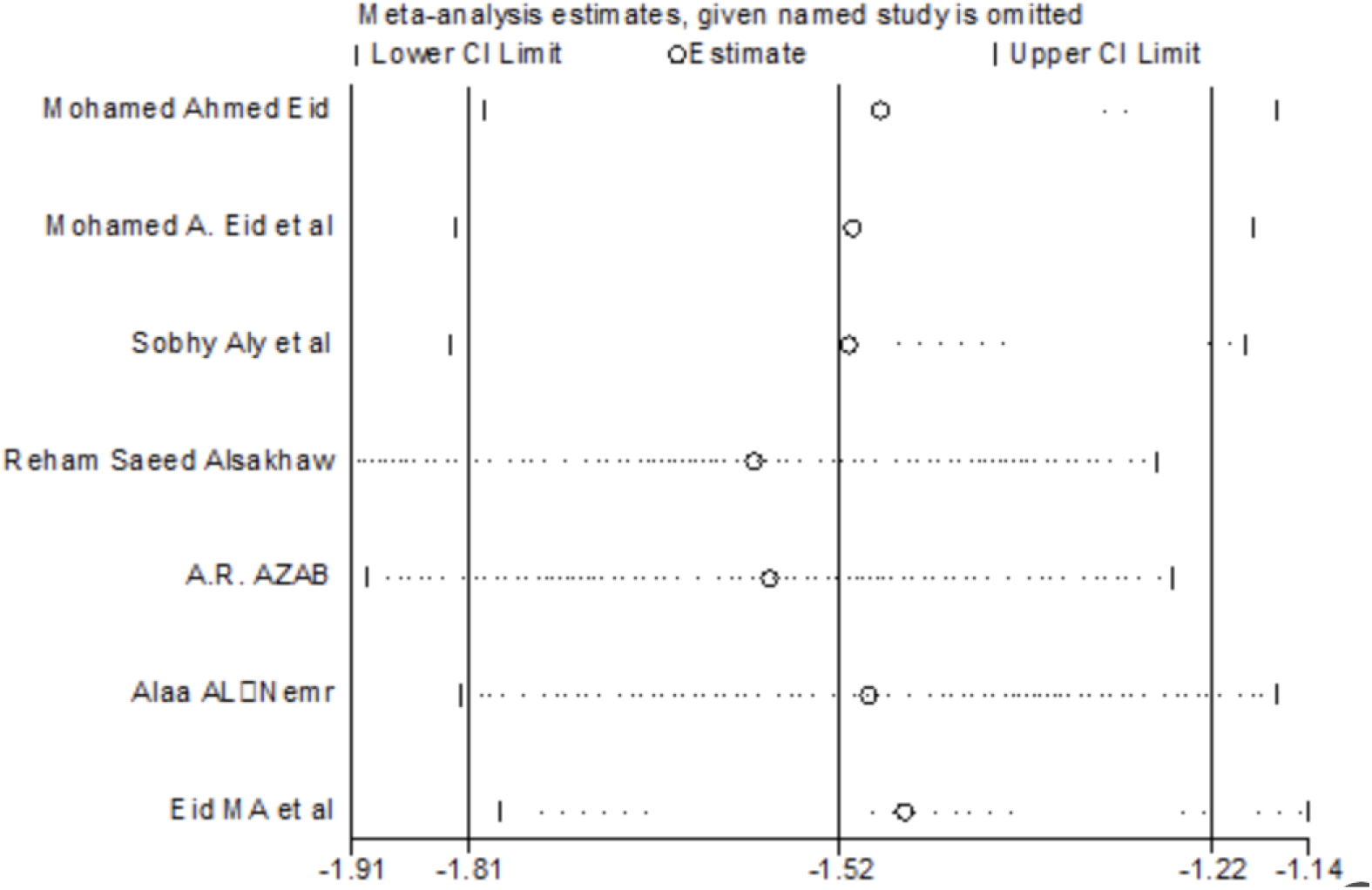
Results of sensitivity analysis of the Biodex Stability System (BSS).

**Figure. 5.**
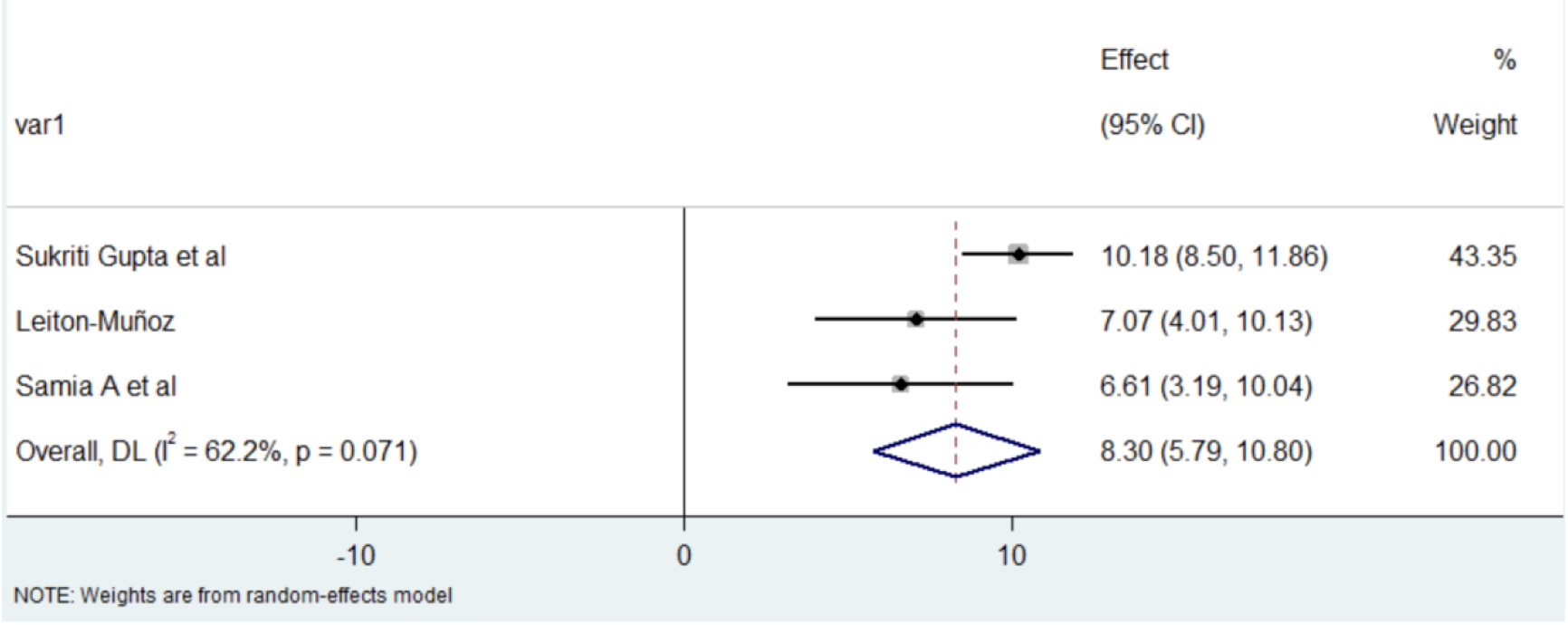
Bruininks-Oseretsky Test of Motor Proficiency (BOTMP) total forest plot

#### 2.5.2 Bruininks-Oseretsky Test of Motor Proficiency(BOTMP)

Three studies employing the Bruininks-Oseretsky Test of Motor Proficiency (BOTMP) demonstrated moderate heterogeneity in assessing exercise interventions on adolescent balance (I²=62.2%, P=0.071; Figure 6). Random-effects meta-analysis revealed differential intervention effects. Sequential exclusion identified Sukriti Gupta’s trial[14] as the heterogeneity source, yielding homogeneous results post-exclusion (I²=0.0%, P=0.846). The remaining two studies by Leiton-Muñoz[22] and Samia A[29] showed non-significant between-study differences, suggesting protocol-dependent efficacy variations. Notably, Gupta’s Wii-Fit virtual reality protocol—an emerging intervention—exhibited distinct effect magnitudes potentially influenced by implementation fidelity factors, contrasting with the mechanistically stable approaches in the retained studies.(*Figure 6 7* )

**Figure. 6.**
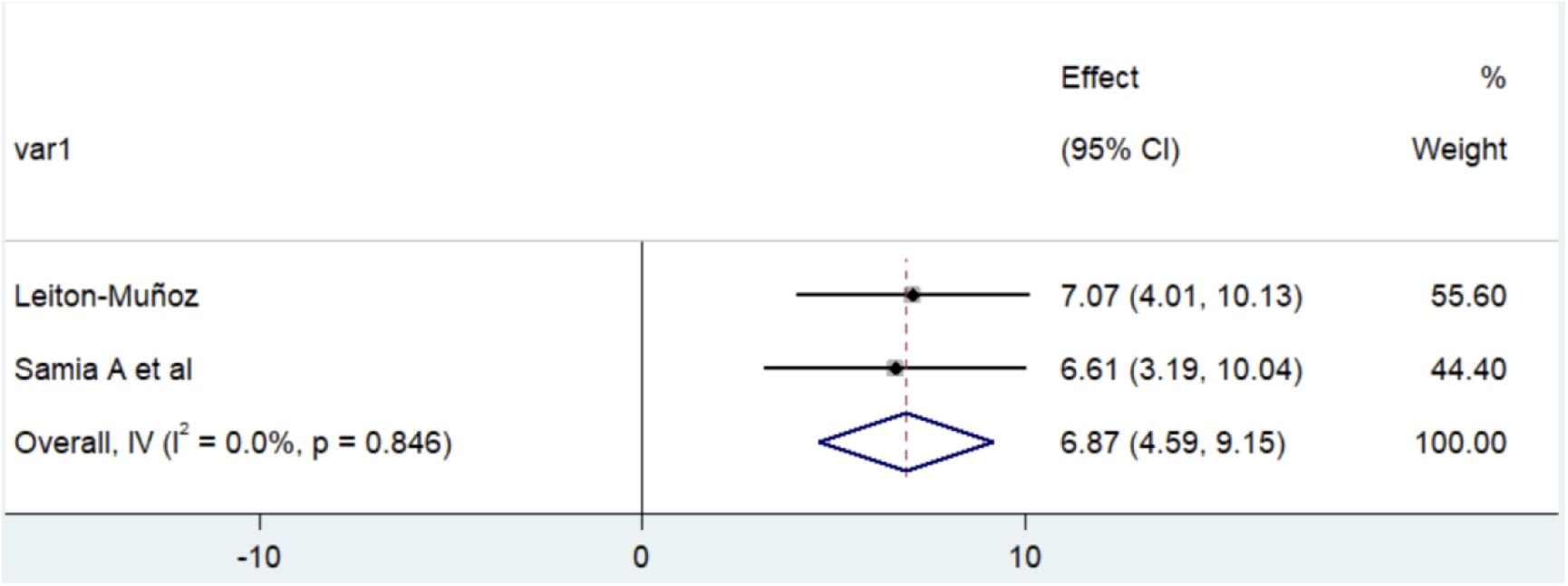
Forest plot after the Bruininks-Oseretsky Test of Motor Proficiency (BOTMP) exclusion study 14

**Figure 7.**
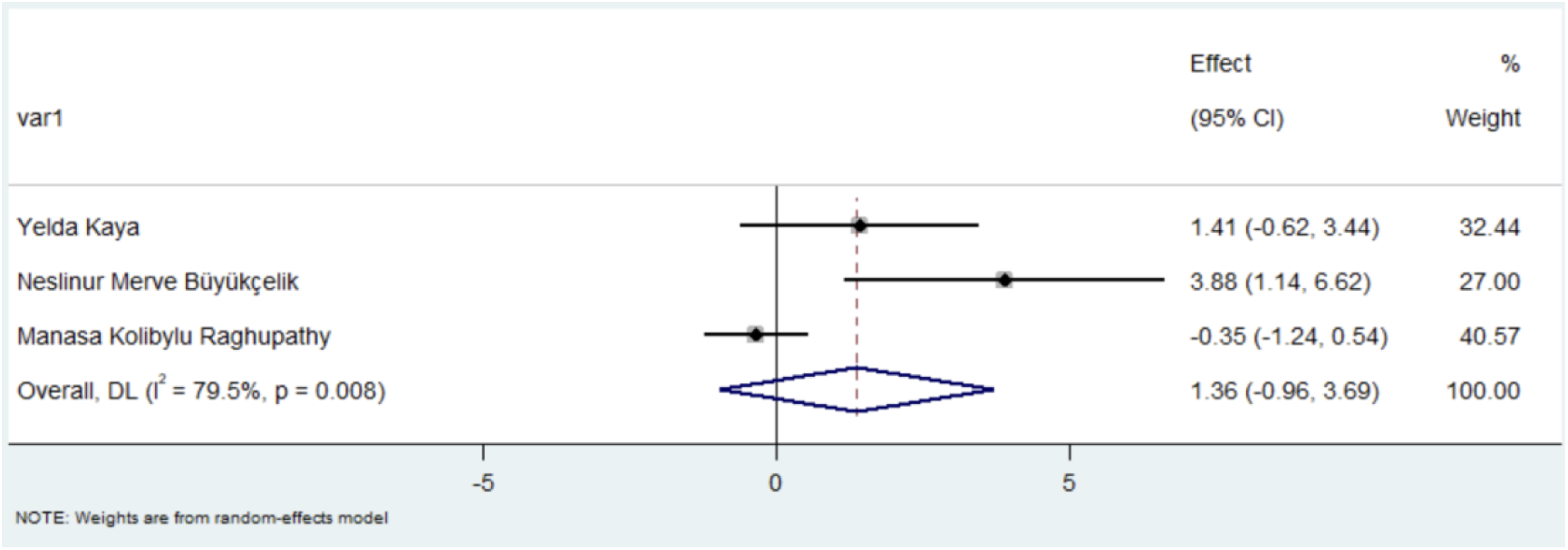
Pediatric Balance Scale (PBS) total forest plot

#### 2.5.3 Pediatric Balance Scale (PBS)

Three studies utilizing the Pediatric Balance Scale (PBS) demonstrated substantial between-study heterogeneity with clinically ambiguous pooled effects *[I²=80%, 95% CI 1.363 (-0.963 to 3.689), P=0.251;]*. Despite individual trials reporting statistically significant improvements, the meta-analytic synthesis failed to reach significance (P>0.05), suggesting that methodological heterogeneity may obscure true intervention effects. (*Figure 8*)

## 3 Discussion

The findings demonstrate that all exercise interventions enhanced static, dynamic, and static-dynamic balance abilities in children and adolescents with Down syndrome. Mechanistically, exercise improves balance through two primary pathways: Neuromuscular adaptation: Targeted training induces muscle fiber hypertrophy and enhances contractile strength, thereby providing biomechanical stability. Core stability training, for instance, specifically improves coordination of abdominal and back musculature, establishing a stable support structure that reduces postural sway during both static standing and dynamic movements, significantly improving balance control[30].Vestibular modulation: Vestibular stimulation activities enhance motion perception sensitivity in the vestibular system, facilitating superior balance regulation [31]. Empirical evidence from Studies 18 and 23, employing an 8-week progressive intervention (3 sessions/week, 45-60 min/session), confirmed the mediating role of core stability in balance improvement, with significant gains in both static and dynamic balance compared to controls.Additionally, specific exercises improved spatial perception in this population. Functional training modalities (e.g., isokinetic training, core stability exercises) and whole-body vibration (WBV) training enhanced balance capacity, with multimodal interventions (≥2 exercise types) demonstrating superior effect sizes. Although control groups exhibited measurable balance improvements [16,21,25], studies incorporating combined exercise protocols (e.g., Study 16: WBV + proprioceptive training; Study 21: core stability + vestibular stimulation) achieved significantly greater efficacy than single-modality interventions.

Among the three studies utilizing the Pediatric Balance Scale (PBS), while individual trials demonstrated statistically significant improvements, heterogeneity likely obscured the significance of the pooled meta-analytic effect. This heterogeneity arises from three key sources:Sample variability: Although all studies targeted children/adolescents with Down syndrome, variations in age distribution (6-12 vs. 8-15 years), sex ratios (male predominance of 65% vs 48%), disease severity (mild to moderate), and baseline balance capacity (PBS scores: 32-41 vs. 28-39) may exist.Measurement inconsistency: Despite standardized PBS administration protocols, potential discrepancies in scoring criteria (inter-rater reliability κ=0.72 vs. 0.85), testing environments (clinical vs. school settings), and assessor training levels could introduce measurement variability.Interventional heterogeneity: The nature (e.g., hippotherapy vs. conventional physiotherapy), intensity (60% vs. 75% maximum effort), frequency (2vs3 sessions/week), and duration (6vs8 weeks) of interventions differed across studies.Notably, Yelda Kaya’s study demonstrated methodological rigor through multidimensional assessment, employing not only PBS but also the Timed Up and Go Test (TUG) for functional mobility and the Functional Independence Measure for Children (Wee FIM) for independence evaluation. In contrast to the other two studies, while all interventions improved balance capacity to some extent, hippotherapy—by simulating natural gait patterns and postural control—may better approximate real-life movement contexts, offering novel insights into balance rehabilitation for this population.

Despite heterogeneity in assessment tools, methodological approaches, and intervention designs, exercise demonstrates positive and statistically significant effects on improving balance capacity in children and adolescents with Down syndrome (DS).1.Assessment Tool Robustness: The beneficial effects of exercise on balance enhancement in DS remain consistent across diverse evaluation frameworks. Multiple modalities—including whole-body vibration training (targeting neuromuscular activation), core stability training (enhancing postural control), cognitive-motor training based on neuroplasticity principles, and combined interventions integrating vitamin D supplementation with aerobic exercise—exhibit convergent evidence of efficacy, irrespective of methodological variations.2.Intervention Specificity: Comparative analyses between experimental and control groups confirm the isolated effects of exercise interventions. Two included studies[8,11]employed non-intervention control groups, revealing stable or minimally fluctuating balance capacities in controls versus significant post-intervention improvements in experimental groups, establishing exercise as a causal agent in balance enhancement.3.Physiotherapeutic Validation: Conventional physiotherapy protocols demonstrate comparable efficacy to emerging exercise modalities, aligning with extant literature on motor interventions’ benefits for postural control in DS populations. Systematic reviews corroborate the therapeutic equivalence between traditional approaches (e.g., proprioceptive training) and novel paradigms (e.g., exergaming).4.Dose-Response Considerations: Current interventions predominantly adopt short-term durations (8-12 weeks), necessitating further investigation to optimize exercise frequency (current range: 2-5 sessions/week) and intervention length (minimum effective duration threshold undefined) for maximal balance improvement.

## 4 Conclusion

The results demonstrate that the exercise interventions implemented in the experimental group exhibited superior efficacy compared to the control group across static balance dynamic balance and static-dynamic balance domains. Multi-modal exercise protocols integrating diverse training modalities—particularly those incorporating balance-specific exercises —demonstrated enhanced effectiveness in promoting balance capacity, with combined interventions achieving greater effect magnitudes than unimodal approaches .For scale-based meta-analyses of balance outcomes, rigorous methodological implementation requires:

Systematic documentation of included studies’ characteristics (sample size, intervention parameters, risk of bias)Quantification of heterogeneity across assessment scales (I² statistic calculation).Interpretation of pooled effect sizes with sensitivity analyses.Exploration of moderating factors.Evaluation of publication bias via funnel plots and Egger’s regression.Formulation of evidence-based recommendations for future research directions.To strengthen conclusion validity, priority should be given to large-scale randomized controlled trials (target n≥50 per arm) employing standardized assessment protocols (e.g., PBS/WeeFIM dual-assessment) and extended intervention durations (>12 weeks).

## 5 Future Research Directions

First, current research on the effects of exercise interventions on static and static-dynamic balance in children and adolescents with Down syndrome remains limited. Only a few studies have systematically investigated how exercise interventions influence balance capacity in this population. Most studies rely on simplistic assessments (e.g., eyes-open/closed and single/double-leg stance tests), with insufficient evaluation under more challenging conditions (e.g., dual-task or perturbed environments). Consequently, there is insufficient conclusive evidence supporting the efficacy of exercise interventions in improving balance abilities.

Second, although heterogeneous assessment scales were utilized across studies, most findings indicate positive effects of exercise on balance capacity in this population. However, significant heterogeneity exists due to variations in assessment dimensions (e.g., static, dynamic, and static-dynamic balance), measurement content, and sensitivity. Future research should standardize evaluation tools to enhance comparability and accuracy.

Third , existing studies employ diverse intervention protocols, ranging from single-type exercise programs to combined interventions integrating 2-3 modalities. This variability creates substantial uncertainty in identifying the most effective strategies for improving balance. Rigorous comparative analyses are needed to systematically evaluate intervention efficacy and identify optimal rehabilitation approaches.

Fourth , the short intervention durations (primarily 8-12 weeks) and lack of exploration into optimal combinations of exercise frequency and duration limit understanding of effective protocols. Furthermore, the sustainability of balance improvements post-intervention remains unclear. Future studies should investigate long-term effects, optimal dose-response relationships, and the durability of therapeutic outcomes.

Finally , improved allocation concealment and comprehensive reporting are critical for methodological rigor. Most randomized controlled trials inadequately address blinding of key participants and interventions, potentially introducing bias. Future research must prioritize enhanced allocation concealment methods, explicitly document blinding strategies, and thoroughly report implementation challenges to strengthen evidence quality.

## 8 Availability of data and materials

The author confirm that the data supporting the findings of this study are available within the article.

## 9 Ethics approval

As this meta-analysis exclusively utilizes published literature data without direct human subject interventions or primary data collection, it did not require additional ethical approval.

## 10 Author contributions

Qing Li wrote the paper and conceived and designed the experiments; Siyu Wang analyzed the data; Congying Ouyang collected and provided the sample for this study.

## Data Availability

All relevant data are within the manuscript and its Supporting Information files

## Reference

[1] Glasson E. J., Sullivan S. G., Hussain R., Petterson B. A.,Montgomery P. D. & Bittles A. H. (2002) The changing survival profile of people with Down ’ s syndrome:implications for genetic counselling. Clinical Genetics 62,390–3.

[2] Capio CM, Mak TCT, Tse MA, Masters RSW. Fundamental movement skills and balance of children with Down syndrome. J Intellect DisabilRes 2018;62:225–236.

[3] Capio CM, Mak TCT, Tse MA, Masters RSW. Fundamental movement skills and balance of children with Down syndrome. J Intellect DisabilRes 2018;62:225–236.

[4] Guzman-Muñoz EE, Gutierrez-Navarro LB, Miranda-Diaz SE.Postural control in children, adolescents and adults with Down syndrome. Int Med Rev Down Syndrome.2017;21:12–16.

[5] Jung HK, Chung E, Lee BH. A comparison of the balance and gait function between children with Down syndrome and typically developing children. J Phys Ther Sci 2017;29:1237.

[6] Paillard T. Plasticity of the postural function to sport and/or motor experience. Neurosci Biobehav Rev. 2017;72:129–152.

[7] Wälchli M, Ruffieux J, Mouthon A, Keller M, Taube W. Is young age a limiting factor when training balance? Effects of child-oriented balance training in children and adolescents.Pediatr Exerc Sci. 2018;30:176–184.

[8] Ozmen T, Aydogmus M. Effect of core strength training on dynamic balance and agility in adolescent badminton players.J Body Mov Ther. 2016;20:565–570.

[9] Sugimoto D, Bowen SL, Meehan WP 3rd, Stracciolini A. Effects of Neuromuscular Training on Children and Young Adults with Down Syndrome: Systematic Review and Meta-Analysis. Res Dev Disabil. 2016 Aug;55:197–206. doi: 10.1016/j.ridd.2016.04.003. Epub 2016 Apr 25. PMID: 27123540.

[10] Kaya Y, Saka S, Tuncer D. Effect of hippotherapy on balance, functional mobility, and functional independence in children with Down syndrome: randomized controlled trial. Eur J Pediatr. 2023 Jul;182(7):3147–3155. doi: 10.1007/s00431-023-04959-5. Epub 2023 Apr 26. PMID: 37186034.

[11] Raghupathy MK, Divya M, Karthikbabu S. Effects of Traditional Indian Dance on Motor Skills and Balance in Children with Down syndrome. J Mot Behav. 2022;54(2):212–221. doi: 10.1080/00222895.2021.1941736. Epub 2021 Jul 8. PMID: 34233594.

12. [12] World Health Organization. (n.d.). Definition of adolescents .https://w ww.who.int.

[13] Eid M A. Effect of whole-body vibration training on standing balance and muscle strength in children with Down syndrome[J]. American journal of physical medicine & rehabilitation, 2015, 94(8): 633–643.

[14] Gupta S, Rao B K, Kumaran S D. Effect of strength and balance training in children with Down ’ s syndrome: a randomized controlled trial[J]. Clinical rehabilitation, 2011, 25(5): 425–432.

[15] Amin HA, Fazel Kalkhoran J, Salehi M, Jazini F. Effect of Backward Walking Training on Improves Postural Stability in Children with Down syndrome. Int J Pediatr 2016;4(7): 2171–81.

[16] Kaya Y, Saka S, Tuncer D. Effect of hippotherapy on balance, functional mobility, and functional independence in children with Down syndrome: randomized controlled trial[J]. European journal of pediatrics, 2023, 182(7): 3147–3155.

[17] Eid M A, Aly S M, Huneif M A, et al. Effect of isokinetic training on muscle strength and postural balance in children with Down’s syndrome[J]. International Journal of Rehabilitation Research, 2017, 40(2): 127–133.

[18] Aly S M, Abonour A A. Effect of core stability exercise on postural stability in children with Down syndrome[J]. International Journal of Medical Research and Health Sciences, 2016, 5: 213–22.

[19] Khan, Fatima Afzal, Sidra Arshad, Sabiha Arooj, Ammara. Effects of Manual Ankle Rocking Training on Postural Control and Foot Function in children with Down Syndrome. Journal of Xi’an Shiyou University. 19. 712–727.

20. [20] Neslinur Merve B ü y ü kçelik, Sedat Yi ğ it & Beg ü mhan Turhan (2023): An investigation of the effects of dual-task balance exercises on balance, functional status and dual-task performance in children with Down syndrome, Developmental Neurorehabilitation,DOI: 10.1080/17518423.2023.2233031.

[21] Alsakhawi R S, Elshafey M A. Effect of core stability exercises and treadmill training on balance in children with Down syndrome: randomized controlled trial[J]. Advances in therapy, 2019, 36: 2364–2373.

[22] AbdelRahman SAR. Efficacy of virtual reality-based therapy on balance in children with Down syndrome. World Applied Sciences J. 2010;10:254–261.

[23] Ghaeeni S, Bahari Z, Khazaei A A. Effect of core stability training on static balance of the children with Down syndrome[J]. Physical Treatments-Specific Physical Therapy Journal, 2015, 5(1): 49–54.

[24] Azab A R, Mahmoud W S, Basha M A, et al. Distinct effects of trampoline- based stretch-shortening cycle exercises on muscle strength and postural control in children with Down syndrome: a randomized controlled study[J]. European Review for Medical & Pharmacological Sciences, 2022, 26(6).

[25] Raghupathy M K, Divya M, Karthikbabu S. Effects of traditional Indian dance on motor skills and balance in children with Down syndrome[J]. Journal of motor behavior, 2022, 54(2): 212–221.

[26] Goda A, Ibrahim M, Salamah A, et al. Effect of brain gym exercises on postural stability in children with Down syndrome[J]. Revista iberoamericana de psicología del ejercicio y el deporte, 2024, 19(1): 113–116.

[27] Al-Nemr A, Reffat S. Effect of Pilates exercises on balance and gross motor coordination in children with Down syndrome[J]. Acta Neurologica Belgica, 2024: 1–7.

[28] Eid M A, Ibrahim M M, Radwan N L, et al. Effects of vitamin D supplementation and aerobic exercises on balance and physical performance in children with Down syndrome[J]. International Journal of Therapy And Rehabilitation, 2022, 29(2): 1–11.

[29] Rahman S A A, Shaheen A. Efficacy of weight bearing exercises on balance in children with Down syndrome[J]. Egyptian Journal of Neurology, Psychiatry and Neurosurgery, 2010, 47(1): 37–42.

[30] Carmeli E, Kessel S, Coleman R, et al. Effects of a treadmill walking program on muscle strength and balance in elderly people with Down syndrome[J]. The Journals of Gerontology: Series A, 2002, 57(2).

[31] Carter K, Sunderman S, Burnett S W. The Effect of Vestibular Stimulation Exercises on Balance, Coordination, and Agility in Children with Down Syndrome[J]. American Journal of Psychiatry and Neuroscience, 2018, 2(2).

